# Artificial Intelligence-Based Chatbots in Genetic Counseling Practice: Current Uptake, Utilization, and Perspectives

**DOI:** 10.64898/2026.05.21.26353789

**Authors:** Nicole Daley, Alyssa Griswold, Laura Moreno, Alyson Evans Floyd, Dat Duong, Benjamin D. Solomon, Rebekah L. Waikel

## Abstract

AI-driven chatbots have been utilized in healthcare to automate administrative tasks, improve patient education, and expand access to medical information; however, their role in genetic counseling remains underexplored. To investigate the adoption, perceptions, and potential utility of AI-based chatbots in genetic counseling practice, 217 genetic counselors and genetic counseling students from across North America were surveyed regarding chatbot usage, confidence in their application, and perceived benefits and limitations. While most participants (166/217; 76.5%) reported using general AI chatbots outside of clinical settings, far fewer (18/204; 8.8%) reported using or recommending clinical genetics chatbots in clinical practice. For those that used clinical genetics chatbots, the primary purpose was for communication with at-risk family members (11/18; 61.1%) and patient education (10/18; 55.6%). Confidence in chatbot technology varied, with highest confidence in gathering family history information (81/199; 40.7%) and lowest confidence in their ability to disclose variants of uncertain significance or positive genetic testing results (5/199; 2.5%). The greatest perceived benefits included reducing repetitive tasks (165/195, 84.6%) and allowing for time for other tasks (141/195; 72.3%), while major concerns revolved around patient comprehension (167/195; 85.6%) and having accurate, up-to-date information (145/195; 74.4%). Despite some concern about AI replacing human counselors, most participants reported they felt there was potential for chatbots to enhance workflow efficiency (128/195; 65.6%) if properly integrated and regulated. Limited AI training was identified as a barrier to adoption (16/195; 8.2% received training), highlighting a need for structured education on AI applications in genetic counseling. These findings suggest that AI chatbots hold promise as supplementary tools, but significant challenges must be addressed before widespread implementation in genetic counseling practice.

## Introduction

Genetic counseling is a rapidly expanding field within healthcare, with a predicted 9% growth from 2024 to 2034^1^. Iincreasing demand for genetic testing and personalized medicine drives this surge. However, the increase in demand for genetic counselors and genetic testing exceeds the availability of trained counselors and support staff, creating a pressing need for changes to the field^2^.

In many genetics clinics, high workload levels and lack of support staff have resulted in longer patient wait times and diminished quality of patient care^3^. According to one study, approximately 25% of genetic counselors’ time is consumed by administrative and repetitive tasks that could be delegated^4^. Tasks such as pretest counseling, patient scheduling, contacting patients before appointments, and disclosing negative or low-risk results do not always require the expertise of a genetic counselor and could be handled by support staff or automated systems^4^. Additionally, 71% of surveyed genetic counselors reported being willing to delegate such tasks^4^. Moreover, many genetic counselors feel they lack adequate support staff, which is a major contributor to burnout, but clinics face difficulties hiring more personnel due to insufficient funding and institutional support^5^. Higher levels of burnout are linked to reduced empathy, lower positive unconditional regard, and a desire to reduce clinical care time^5^.

Digital tools, such as telehealth platforms and electronic health records, have demonstrated utility in the field of genetic counseling by facilitating communication and reducing administrative burdens^6^. Artificial intelligence (AI) can assist in various aspects of patient care, from risk assessment to personalized treatment plans^6^. Previous studies have illustrated that patients and providers have generally positive experiences with AI in medical settings, but demographic and other factors can shape a preference for more traditional clinic formats^7^.

Chatbots, a form of AI that use algorithms to interpret and simulate human conversation, are currently utilized in various healthcare settings to provide information, schedule appointments, and other tasks like offering mental health support^6^. In genetics clinics, chatbots are beginning to be used for tasks such as patient intake and follow-up communication^8^. Studies thus far illustrate that genetic counselors, other medical professionals, and patients have found chatbots to be a promising way to provide accurate information about genetics, increase genetic counseling accessibility, and enhance clinical productivity, yet they highlight concerns that the technology may not be optimal for certain tasks^8^. Additionally, studies have also demonstrated the feasibility of chatbot use in specific genetics contexts, including pretest genetic services and hereditary cancer result disclosure, while simultaneously highlighting ongoing concerns regarding chatbot accuracy and the need for substantial oversight, iterative refinement, and validation prior to clinical implementation^9,10^. Recent patient-centered studies found that chatbot-supported pretest genetic services may promote patient engagement and influence genetic testing decision-making, suggesting a potential role for chatbots in improving access to genetics care ^11^.

Existing research suggests the potential of AI to streamline and improve efficiency in genetics clinics; however, the potential role of chatbots and the perspective of genetic counselors on their utility in genetics clinics has not been extensively studied. This study builds upon this background by evaluating the feasibility and utility of integrating chatbots into genetic counseling practice, as well as exploring the potential of this technology to enhance the efficiency of genetic counseling. The aim of this study was to identify the level of confidence in and the uptake of AI-based chatbots by genetic counselors. The study team assessed the uptake of clinical genetics chatbots in genetics clinics, evaluated perspectives on trust in clinical genetics chatbots, and identified strengths and limitations of current clinical genetics chatbot technology.

## Materials and Methods

### Participants Recruitment

Board-certified/board-eligible genetic counselors (n=1065) and genetic counseling students (n=310) were contacted directly via email. Contact information for potential participants was obtained via professional networks and online sources. Survey data were collected from October 2024 to April 2025. Only surveys from individuals who completed at least the second section, which included questions about general chatbot usage, were considered participants and included in the analysis.

The study was approved by the Bay Path University Institutional Review Board (IRB# 2024_Daley/Griswold).

### Survey Instrument

An online survey was designed in Qualtrics (Provo, Utah, United States); the survey included four sections: demographic questions (section 1), general chatbot tutorial and questions on familiarity and use (section 2), healthcare-specific chatbot tutorial and questions on usage and perceived clinical utility (section 3), perception questions about the benefits and limitations of chatbots (section 4). Question types included Likert scale questions (4-point confidence scale, e.g., 1 = not confident, 4= highly confident), multiple choice, and open-ended questions. The survey was developed de novo (no relevant validated instruments exist) based on previous work^3,4,12,13^ by a team of two genetic counseling students, three genetic counselors, a medical geneticist, and a computer scientist. Survey participants were able to proceed through the survey without needing to respond to each question before moving to the next question. Therefore, the number of responses per question varies from the total number of participants. The survey can be found at: https://github.com/rlwaikel/GCandChatbots.

### Data Analysis

Survey data were exported from Qualtrics and stored and analyzed using Microsoft Excel. Participants were categorized into two groups: genetic counselors; genetic counseling students. Descriptive statistics were used to analyze demographic data. Frequencies were employed to analyze trends and patterns in the multiple-choice questions. For Likert scale questions, the average confidence rating was calculated to gauge participants’ general confidence levels in different scenarios. To measure the association among the responses for any two questions of interest (e.g., “general chatbot use” and “clinical genetics chatbot use”), we used the Chi-square test of independence. For certain analyses within the study, responses from genetic counseling students were excluded and results from practicing genetic counselors were analyzed related to questions that focused specifically on the clinical use of chatbots, an area in which students do not yet have direct experience. Open-ended responses were manually coded and analyzed using content analysis.

### Participant demographics

A total of 217 genetic counselors and genetic counseling students (n=160 genetic counselors, response rate=15.0%; n=57 genetic counseling students, response rate=18.4%) participated in the survey. Participants included genetic counselors and students from 38 different US states and 3 different provinces in Canada. Most genetic counselors reported their primary place of employment as an academic medical or research center (99/160, 61.9%) and primarily working in a clinical setting (117/160; 73.1%). Over one-quarter of genetic counselors reported currently working in multiple specialties (44/160; 27.5%); the most reported specialties were cancer (59/160 36.9%) and pediatrics (35/160; 21.9%). Genetic counselor participants represented both junior and senior genetic counselors with over 35% (56/160) of participants reporting over 10 years of practice experience. See Table 1 and Supplemental Table 1 for additional demographic information.

**Table 1.** Participant Demographics.

|  |  | Frequency | Percent |
| --- | --- | --- | --- |
| Participants<br>(n=217) | Genetic counselors | 160 | 73.7 |
|  | Genetic counseling students | 57 | 26.3 |
| Genetic Counselors<br>Current Area(s) of Practice*<br>(n=160) | Cancer | 59 | 36.9 |
|  | Pediatrics | 35 | 21.9 |
|  | Prenatal | 29 | 18.1 |
|  | General | 28 | 17.5 |
|  | Research | 26 | 16.3 |
|  | Cardiovascular | 17 | 10.6 |
|  | Neurology | 13 | 8.1 |
|  | Laboratory | 11 | 6.9 |
|  | Ophthalmology | 7 | 4.4 |
|  | Other (specialty disease clinics, etc.) | 25 | 15.6 |
| Genetic Counselors<br>Institution Type<br>(n=160) | Academic medical or research center | 99 | 61.9 |
|  | Community based hospital | 39 | 24.4 |
|  | Molecular diagnostic company | 3 | 1.9 |
|  | Other (government, biotech, etc.) | 19 | 11.9 |
| Genetic Counselor<br>Role(s)*<br>(n=160) | Clinical practice | 117 | 73.1 |
|  | Clinical research | 32 | 20.0 |
|  | Laboratory | 13 | 8.1 |
|  | Basic science research | 5 | 3.1 |
|  | Other (administration, teaching, etc.) | 24 | 15.0 |
| Genetic Counselors<br>Years of Practice<br>(n=160) | Less than 1 year | 20 | 12.5 |
|  | 1 to 5 years | 45 | 28.1 |
|  | 5 to 10 years | 39 | 24.4 |
|  | Over 10 years | 56 | 35.0 |

\*Participants could select multiple practice areas and roles.

## Results

### General Chatbot Use

Most participants (166/217, 76.5%) reported prior use of general AI chatbots, with higher uptake reported by students (86.0%; 49/57) than genetic counselors (73.1%; 117/160) (Table 2). The most used chatbots were ChatGPT (155/166, 93.4%) and Google Gemini (42/166, 25.3%) (Supplemental Figure 1). The primary applications of general chatbots for personal use outside clinical practice included: general questions (n=128/166, 75.3%), composing text (n=101/166, 59.4%), and entertainment (62/166, 36.5%) (Supplemental Table 2). Among genetic counselors who reported chatbot use, 23.9% (28/117) reported using general chatbots outside clinical practice at least once a week, compared to more than half of students (25/49; 51.0%) (Table 2).

**Table 2.** Uptake and Uses of General Chatbots Outside and Inside Clinical Practice.

|  |  | Frequency | Percent |
| --- | --- | --- | --- |
| General Chatbot Uptake | All participants (n=217) | 166 | 76.5 |
|  | Genetic counselors (n=160) | 117 | 73.1 |
|  | Students (n=57) | 49 | 86.0 |
| Frequent Users of General Chatbots Outside of Clinical Practice (weekly/daily) | All general chatbot users (n=166) | 53 | 31.9 |
|  | Genetic counselors (n=117) | 28 | 23.9 |
|  | Students (n=49) | 25 | 51.0 |
| General Chatbots Uptake in Clinical Practice | All participants (n=217) | 79 | 36.4 |
|  | Genetic counselors (n=160) | 51 | 31.9 |
|  | Students (n=57) | 28 | 49.1 |
| Frequent Users of General Chatbots Inside of Clinical Practice (weekly/daily) | All participant clinical users (n=79) | 43 | 54.4 |
|  | Genetic counselors (n=51) | 26 | 51.0 |
|  | Students (n=28) | 17 | 60.7 |
| General Chatbot Uses in Clinical Practice* (n=166) | Composing Text | 64 | 38.6 |
|  | Medical related education | 45 | 27.1 |
|  | Scientific research projects | 17 | 10.2 |
|  | Technology support | 16 | 9.6 |
|  | Possible patient diagnoses | 16 | 9.6 |
|  | Diagnostic testing procedures | 5 | 3.0 |
|  | Composing computer code | 4 | 2.4 |
|  | Patient Management | 3 | 1.8 |
|  | Only use outside of clinic, not for clinical | 87 | 52.4 |

\*Participants could report multiple responses.

In a clinical setting, 36.4% (79/217) of total participants reported using general chatbots in their clinical work, with genetic counselors (51/160; 31.9%) reporting lower uptake levels than students (28/57; 38.6%). Among general chatbot users, primary uses in the clinic included composing text (64/166; 38.6%) and medical related education (45/166, 27.1%) (Table 2 and Supplemental Table 2). Greater than half of general chatbot users in the clinic reported using them at least once a week as part of their clinical practice, including both genetic counselors (26/51; 51.0 %) and students (17/28; 60.7%) (Table 2). When asked to rate their knowledge of the use of general chatbots in healthcare, the majority of genetic counselors 60.9% (95/156) reported little to no knowledge, compared to 48.1% (26/54) of students (Supplemental Table 3). A Chi-square of association test revealed a highly significant association between frequent general chatbot personal use and clinical use (at least once a week (χ^2^=81.5, df=1, p<0.001), suggesting that genetic counselors who frequently used chatbots outside of clinic are significantly more likely to use them in clinical settings. Additionally, there was a strong association between frequent general chatbot clinical use and a higher knowledge rating for use of general chatbots in healthcare (χ^2^=23.20, df=1, p<0.001).

### Clinical Genetics Chatbot Use

Regarding the use of domain-specific clinical genetics chatbots, 18 of the 204 (8.8%) participants who completed this section reported either using and/or recommending these chatbots to patients, with higher uptake for genetic counselors (16/153; 10.5%) compared to students (2/51; 3.9%) (Table 3 and Supplemental Table 4). The most used chatbot was Genetic information assistant (Gia®) by Invitae (16/18, 88.9%). The most common reported purpose in clinical practice was “informing at-risk family members of a patient’s test results” (11/18; 61.1%), followed by “education and general information tool for patients” (10/18; 55.6%) and “screening tool to identify potential patients based on family history” (9/18; 50.0%) (Table 3). When genetic counselors who do not use clinical genetics chatbots were asked about their reasons, the most frequent responses were “the technology was not available” (56/135, 41.5%), “doesn’t work with the current clinic workflow” (56/135, 41.5%), and “healthcare facility limits the use of outside technologies” (44/135; 32.6%) (Supplemental Table 5).

**Table 3.**
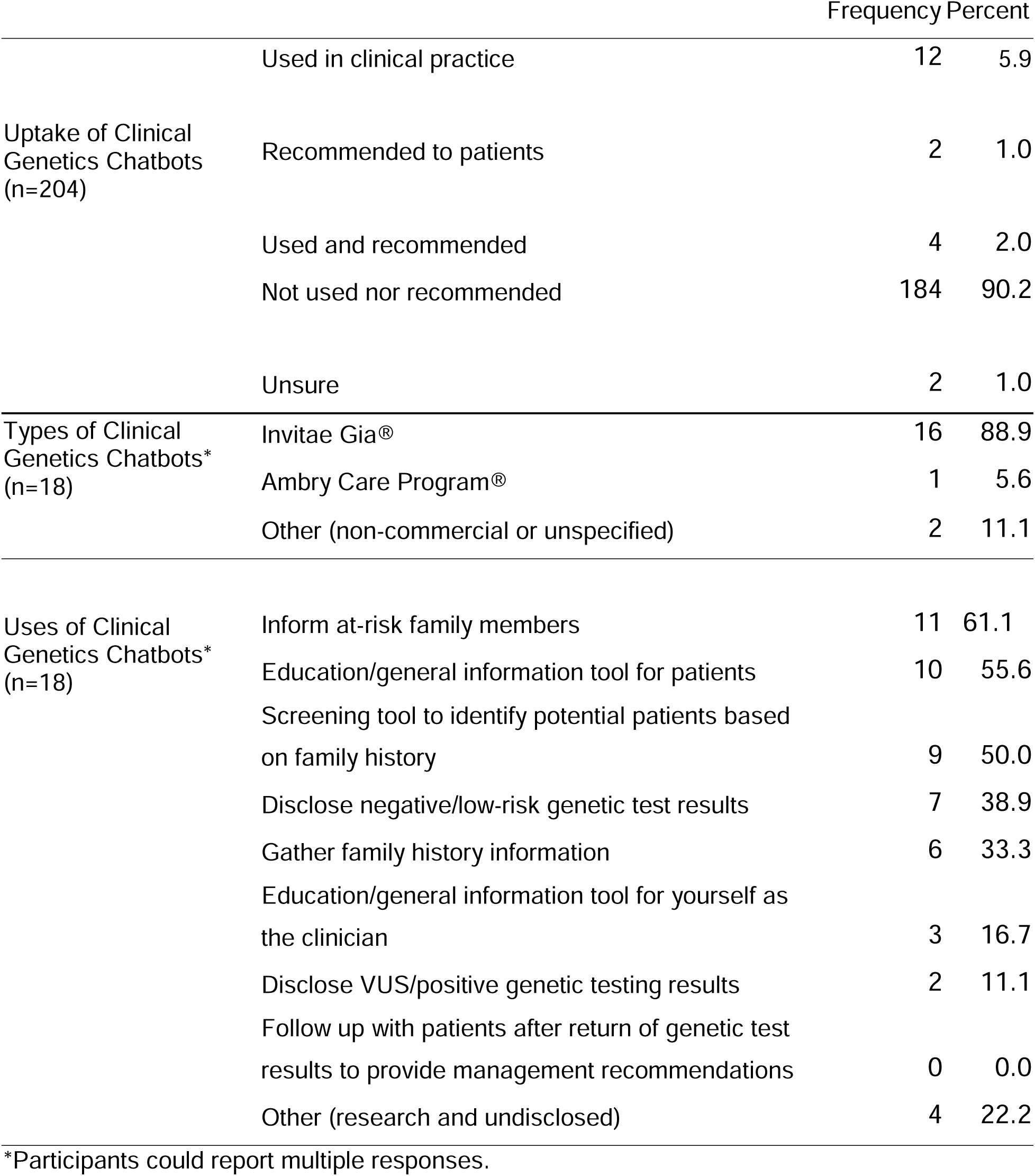
Uptake and Uses of Clinical Genetics Chatbots.

### Confidence in Clinical Genetics Chatbots Technology

To better understand the uptake of clinical genetics chatbots, we asked participants to rate their confidence in the ability of chatbots to perform clinical genetics tasks on a 4-point Likert scale. Chatbot task confidence was highest (greatest percent of participants reporting being highly confident or confident) for gathering family history information (81/199; 40.7%), as an education/general information tool for patients (76/199; 38.2%), and as a screening tool (68/199; 34.2%) (Figure 1). Conversely, confidence was lowest (lowest percentage of participants reporting being confident or highly confident) for tasks such as to disclose variants of uncertain significance or positive genetic testing results (5/199; 2.5%) and to follow-up to provide management recommendations (24/199; 12.1%) (Figure 1). Additionally, when asked if perceived patient comprehension of genetic information is the same with a clinical genetics chatbot as with a human genetic counselor, most respondents (145/199, 72.9%) selected that comprehension is better with a human genetic counselor (Supplemental Table 6). There was a strong association between clinical genetics chatbot use and confidence in clinical genetic chatbot tasks (χ^2^=7.04, df=1, p=0.008) with 43.8% (7/16) of clinical genetics chatbot users reporting confidence in at least half of the eight tasks compared to 16.7% (30/180) of non-users.

**Figure 1.**
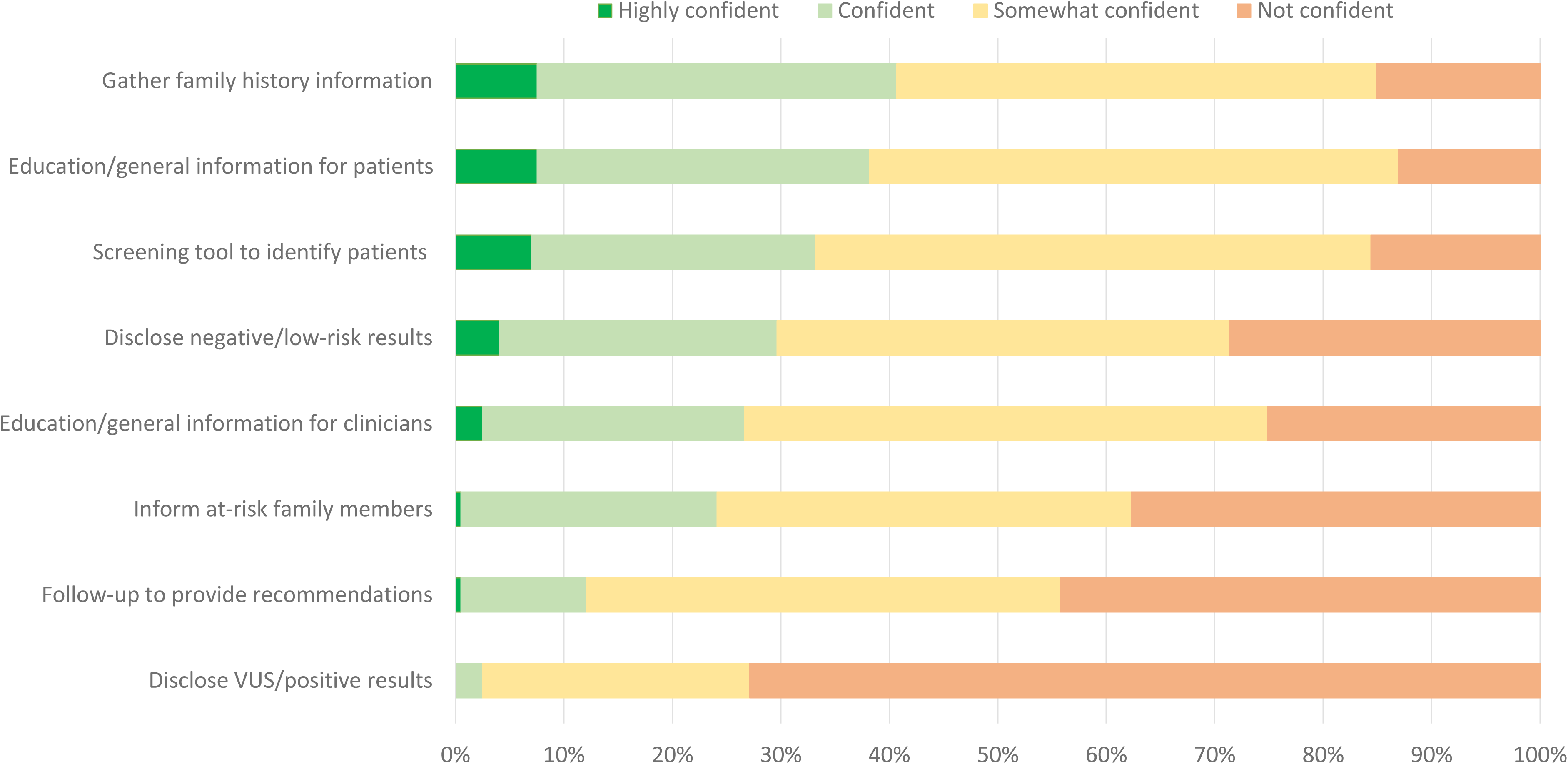
Confidence in Clinical Genetics Chatbot Tasks. Participants (n=199) reported their confidence level on a 4-point Likert scale for clinical chatbot tasks. Percent of total participants are displayed for each task by confidence level. VUS – variant of unknown significance.

### Perceived Benefits and Limitations of Clinical Genetics Chatbots

The greatest perceived benefit for clinicians who use clinical genetics chatbots was the reduction of repetitive tasks (165/195, 84.6%) (Figure 2). Additionally, more than half of participants also perceive that the use of clinical chatbots can allow for time for other tasks (141/195; 72.3%), improve clinic efficiency (128/195; 65.6%), and decrease provider burnout (99/195; 50.8%) (Figure 2). The greatest perceived benefit for patients was ease in reporting family history information (171/195; 87.7%), followed by increased accessibility to information (156/195; 80.0%) (Figure 3 and Supplemental Table 7). Some participants reported no perceived benefits for clinicians (7/195; 3.6%) and patients (8/195; 4.1%) (Figures 2 and 3).

**Figure 2.**
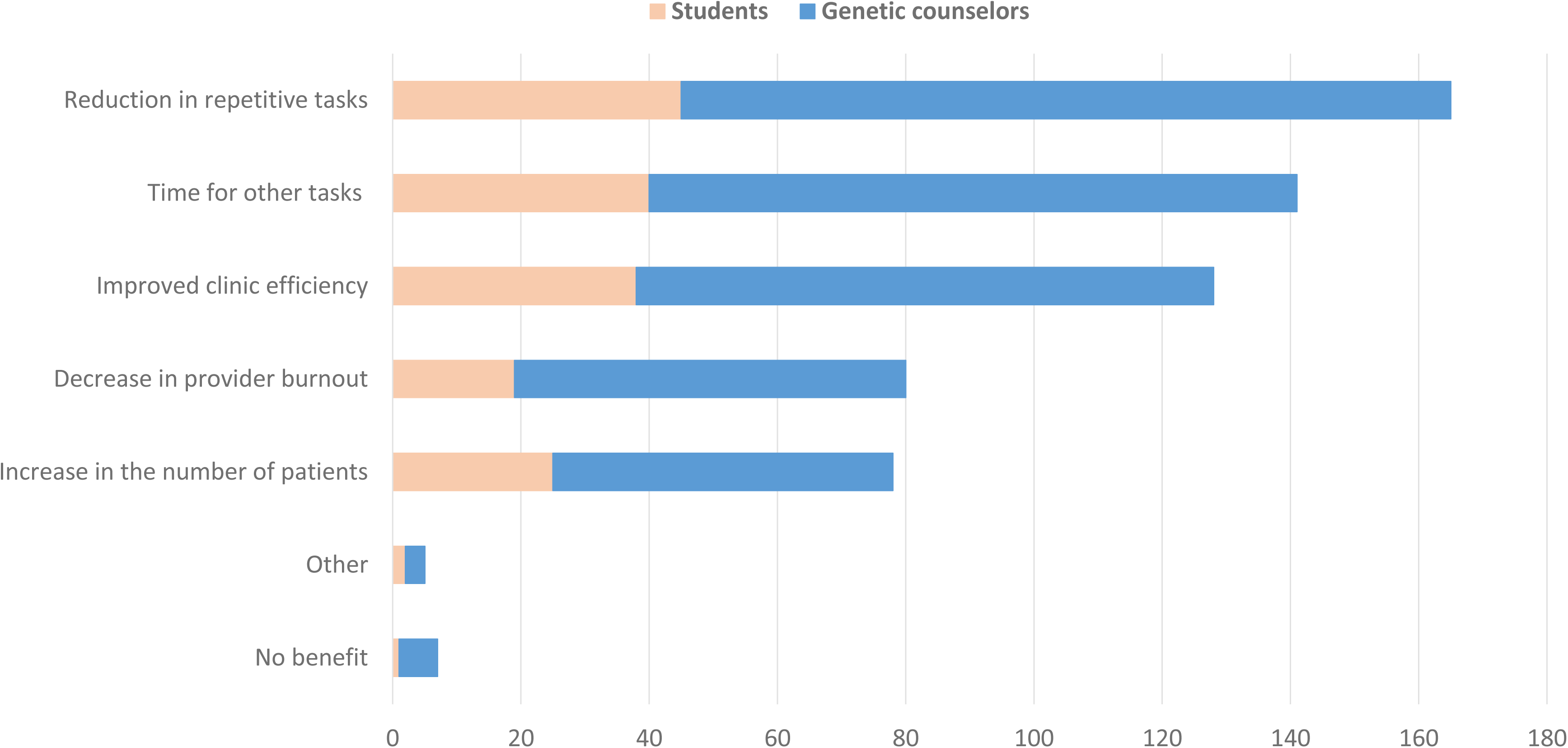
Perceived Benefits of Clinical Genetics Chatbots for Providers. Participants were asked to select from a list of possible benefits for clinical genetic providers, as well as write in responses. Selection titles have been shortened to ease of display. Multiple answers could be selected or written for the 195 participants who completed this section: students (n=50) and genetic counselors (n=145).

**Figure 3.**
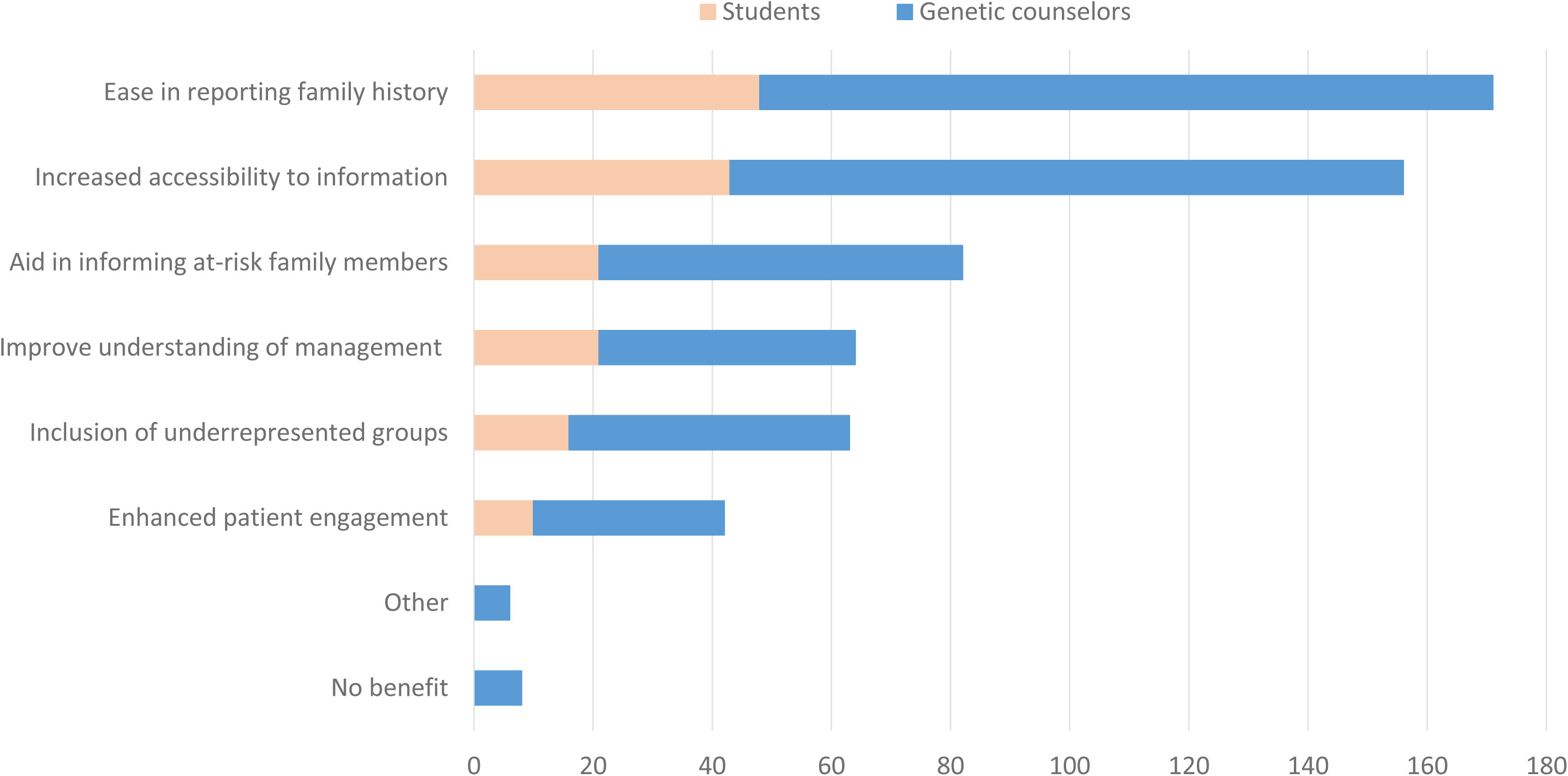
Perceived Benefits of Clinical Genetics Chatbots for Patients. Participants were asked to select from a list of possible benefits for patients, as well as write in responses. Selection titles have been shortened to ease of display. Multiple answers could be selected or written for this section: students (n=50) and genetic counselors (n=145).

When asked about concerns for integrating clinical genetics chatbots into clinical practice, all participants selected at least one concern, with the most frequent responses being inability to assess patients’ understanding of the information provided by the chatbot (167/195; 85.6%) and the chatbots’ ability to provide accurate and up-to-date information (145/195; 74.4%) (Figure 4 and Supplemental Table 8). Many respondents were also concerned about chatbots’ ability to provide content in languages other than English (121/195; 62.1%), the chatbot being mistaken for the genetic counselor or provider (118/195, 60.5%), and chatbots’ compliance with hospital legal and ethical guidelines (115/195, 59.0%) (Figure 4).

**Figure 4.**
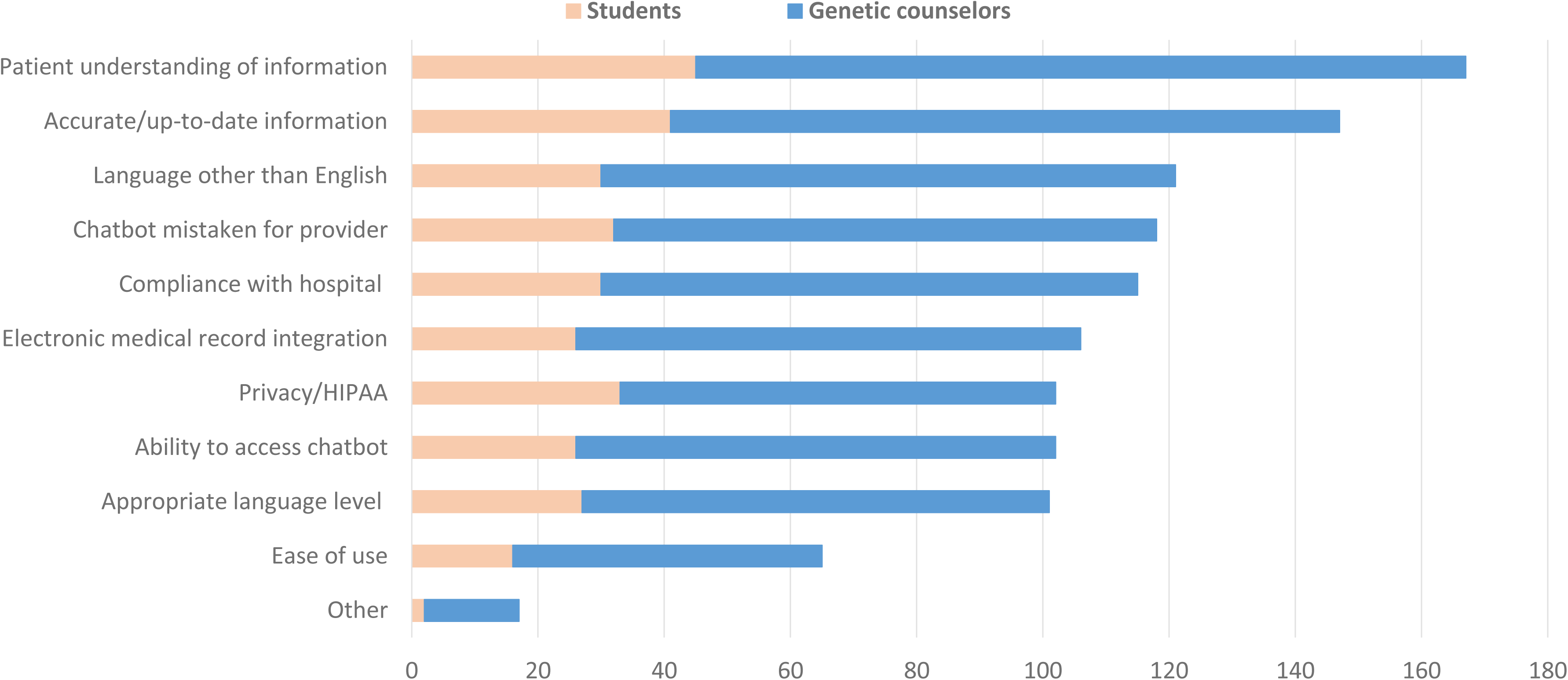
Concerns for Patient Use of Clinical Genetics Chatbots. Participants were asked to select from a list of possible concerns, as well as write in responses. Selection titles have been shortened to ease of display. Multiple answers could be selected or written for this section: students (n=50) and genetic counselors (n=145). HIPPA – Health Insurance Portability and Accountability Act.

When asked about the importance of maintaining the “human element” in genetic counseling, most participants indicated that it is very important (158/195, 81.0%) (Table 4). Participants indicated some concern about AI replacing human counselors, with 47.2% (92/195) indicating some concern, with an additional 30.8% (60/195) indicating moderate or great concern and 22.1% (43/195) indicating no concern. Participants were mostly unsure (n=83/195, 42.6%) of the impact clinical genetics chatbots would have on patient’s perception of the genetic counseling process, with 13.3% (26/195) indicating a positive impact and 11.8% (23/195) indicating a negative impact; the remainder indicated a neutral impact (Table 4). Despite being unsure about impact, participants still indicated an interest in incorporating chatbots: highly interested (20/195; 10.3%), interested (58/195; 29.7%), or some interest (84/195; 43.1%) (Table 4).

**Table 4.** Perceptions and Opinions of Clinical Genetics Chatbots in Genetic Counseling.

|  |  | Frequency | Percent |
| --- | --- | --- | --- |
| Incorporating Clinical Genetics Chatbots into Practice<br>(n=195) | Highly interested | 20 | 10.3 |
|  | Interested | 58 | 29.7 |
|  | Some interest | 84 | 43.1 |
|  | No interest | 33 | 16.9 |
| Perceived Impact on Patients<br>(n=195) | Positive impact | 26 | 13.3 |
|  | Neutral impact | 63 | 32.3 |
|  | Negative impact | 23 | 11.8 |
|  | Unsure | 83 | 42.6 |
| Chatbots Replacing Human Counselors<br>(n=195) | Great concern | 19 | 9.7 |
|  | Moderate concern | 41 | 21.0 |
|  | Some concern | 92 | 47.2 |
|  | No concern | 43 | 22.1 |
| Maintain a Human Element in Interactions<br>(n=195) | Very important | 158 | 81.0 |
|  | Moderately important | 33 | 16.9 |
|  | Slightly important | 4 | 2.1 |
|  | Not important | 0 | 0.0 |

### Chatbot Training

Few participants have received training on clinical genetics chatbots (16/195; 8.2%). Participants report receiving training in graduate school or professional courses (n=8), workshops or conferences (n=4), molecular diagnostics company representative (n=5), and research initiatives (n=4) (Table 5). When asked if genetic counseling programs should incorporate the use of clinical genetics chatbots into their curriculum, 34.9% (68/195) of participants responded that clinical genetics chatbots should be part of the curriculum, whereas nearly half (95/195; 48.7%) indicated that it should be incorporated in the future but not now (Table 5 and Supplemental Figure 2). The most requested clinical genetics chatbot trainings were guidelines on ethical use (n=156/195; 80.0%), training on interpreting chatbot-generated data (145/195; 74.4%), and technical training (n=143/195; 73.3%) (Table 5 and Supplemental Table 9).

**Table 5.** Clinical Genetics Chatbot Training.

|  |  | Frequency | Percent |
| --- | --- | --- | --- |
| Received Training<br>(n=195) | Yes | 16 | 8.2 |
|  | No | 179 | 91.8 |
| Setting for Training*<br>(n=16) | Graduate/professional school course | 8 | 50.0 |
|  | Workshop/conference | 4 | 25.0 |
|  | Molecular diagnostic company | 5 | 31.2 |
|  | Other (research project, etc.) | 4 | 25.0 |
| Training Requested*<br>(n=195) | Guidelines on ethical use | 156 | 80.0 |
|  | Interpreting chatbot-generated data | 145 | 74.4 |
|  | Technical training on using the chatbot | 143 | 73.3 |
|  | Support for troubleshooting | 112 | 57.4 |
|  | Other | 13 | 6.7 |
| Addition of Chatbot Training<br>to Genetic Counseling<br>Curriculum<br>(n=195) | Yes | 68 | 34.9 |
|  | No | 10 | 5.1 |
|  | Maybe in future | 95 | 48.7 |
|  | Unsure | 22 | 11.3 |

Note: Percentages are given based on total number of participants.
\*Participants could provide multiple answers for training setting and types of training requested.

## Discussion

This study aimed to gather information about the use of chatbots in clinical genetics practice. The findings offer insights into the current use, perceptions, and potential future applications of both general and clinical genetics chatbots among genetic counselors and genetic counseling students. The results indicate that while general AI chatbots are widely used in non-clinical settings, their integration into clinical genetics practice remains limited. Furthermore, domain-specific clinical genetics chatbots have been adopted by a small fraction of participants, suggesting barriers to their implementation and acceptance.

### Adoption and Utilization of AI Chatbots

The majority (76.5%) of respondents reported using general chatbots outside of clinical practice, primarily for general inquiries, text composition, and entertainment. Even when chatbots were used within clinical practice, they were primarily employed for composing text and personal education rather than for direct patient interactions. These findings align with previous research, which has shown uneven adoption of AI tools in genetic counseling^12^. While certain AI-driven technologies, such as machine learning models for variant classification and facial analysis tools for dysmorphology assessment, have been widely adopted, AI-driven clinical support tools, including chatbots, remain in the early stages of integration^14^. Our study found an association between general and clinical use through chi square test, suggesting that familiarity with chatbots in non-clinical settings may influence a provider’s willingness to incorporate them into professional practice. However, the overall low adoption rate in clinical settings may indicate significant hesitation or barriers to implementation, as reflected in the limitations identified by respondents when asked about challenges.

### Confidence and Trust in AI for Clinical Genetics

Confidence in chatbots’ ability to perform clinical genetics tasks was highly variable. The highest confidence levels observed were for gathering family history information (40.7%) and patient education (38.2%). Confidence was low for more nuanced tasks such as the disclosure of genetic testing results, particularly VUS or positive findings, with only 2.5% of participants expressing trust in chatbot capabilities for these functions. This suggests that while chatbots may serve as useful supplements for rote tasks in genetic counseling, they are not currently perceived as reliable for delivering complex or sensitive genetic information.

### Perceived Benefits and Barriers to Implementation

Previous research highlights the significant administrative burden on genetic counselors, which consumes a substantial portion of their time and contributes to burnout^5^. A majority of participants in this study expressed confidence that chatbots could save time in clinical practice and allow enhanced productivity for tasks such as research, education, and complex patient interactions. These findings suggest that by optimizing clinic efficiency, chatbots have the potential to alleviate burnout in genetic counseling.

Despite these potential benefits, multiple barriers to adoption were identified. The most frequently cited concern (85.6%) was the chatbot’s inability to assess patient comprehension. This is a fundamental aspect of genetic counseling and extends beyond information delivery to ensure patients fully understand genetic concepts and their health implications. Unlike traditional counseling, where providers can ask clarifying questions, address misunderstandings, and tailor explanations in real time, respondents are unsure if chatbots have valid interactive feedback mechanisms to gauge patient understanding and/ adjust their responses accordingly.

Data from this study also underscored concerns about chatbot accuracy, further highlighting trust-related hesitations regarding the reliability of AI-generated content in clinical settings. Several free responses from participants cited a lack of trust in chatbots; one participant said, “Unless it earns a degree, performs requisite clinical rotations, and passes a board exam, I will not trust a chatbot as I would trust myself or a colleague” (Supplemental Table 10). These issues reflect broader regulatory and ethical concerns documented in previous research, including unresolved questions about AI accuracy, accountability, and patient consent^14^. Many genetic counselors (41.5%) noted that suitable chatbot technology is not available in their practice, while 32.6% reported integration challenges with existing workflows. These concerns are warranted, as previous research found that designing and integrating a breast and ovarian cancer specific chatbot into clinical practice required major iterations to refine the chatbot content, improve usability, and reduce fallback or wrong answers to <15% error^9^. Similarly, Coen et al. demonstrated that substantial prompt engineering and iterative refinement were necessary to improve chatbot performance for hereditary cancer result disclosure, further emphasizing the challenges of ensuring accuracy and reliability in clinical genetics chatbot applications^10^. Lastly, Siglen et al. found that while chatbots can improve access to reliable genetic information, the results demonstrate many of the same concerns identified in our research—namely, that technological, logistical, and ethical challenges may hinder institutional support for chatbot integration^9^. Free-response data reinforced these themes, with participants expressing uncertainty about how to incorporate chatbots into their practice or where to begin implementation. These findings about the benefits and limitations of clinical genetics chatbots suggest that while there is general openness and desire for chatbot-assisted clinical practice, concerns regarding accuracy, security, and ease of integration need to be addressed before widespread adoption can occur.

### Impact on Genetic Counseling and Patient Perception

There is strong agreement among genetic counselors and students about the importance of preserving the human element in genetic counseling. Given the interpersonal nature of the profession, it is not surprising that most respondents did not believe that their role could become obsolete or fully replaced by technology. However, data from this study indicates that participants are still concerned about the potential for AI to replace genetic counselors. These findings parallel concerns raised by Gordon et al., who observed that some genetic counselors worry technology could diminish the demand for their services, particularly if other healthcare providers or patients themselves begin using digital tools to independently deliver or access genetic counseling services^15^. However, this concern contrasts with findings from our study, in which most respondents viewed chatbots and AI tools as valuable adjuncts rather than direct substitutes for human professionals. Additionally, recent research by Yi et al. evaluating chatbot-supported pretest genetic services found that patient interactions with chatbots were associated with engagement in genetic testing and decision-making^11^. These findings align with participants’ perceptions in our study, suggesting that chatbots may improve access to genetics services and support aspects of patient education and screening. Nevertheless, many respondents expressed uncertainty regarding the impact of chatbots on patient perceptions of genetic counseling, highlighting an area where further research is needed, particularly around patient experience and satisfaction with the technology in genetic counseling services.

### Education and Training Needs

There is a significant lack of formal AI training among genetic counseling professionals, with only 8.2% of participants reporting having received AI-specific education. This knowledge gap is a critical issue, as one of the primary barriers to chatbot adoption in genetic counseling appears to be discomfort with technology rather than outright opposition to its use. One participant noted:

> “As someone who works in a clinic with a 6+ month long wait list, I think we as providers have to set aside some of our fear and discomfort and embrace technology that can help reduce wait times and get more patients the services they need” (Supplemental Table 10).

This finding highlights a general hesitancy rooted in unfamiliarity rather than skepticism about the potential benefits of chatbots. Most participants expressed support for incorporating AI and chatbot training into genetic counseling programs. However, many felt that such integration should be postponed until the technology is further refined and its applications are better understood. This presents a paradox: technological advancements in AI for genetic counseling are unlikely to accelerate unless they are more widely integrated into practice, yet widespread integration is difficult to achieve without proper education and training. Without structured educational initiatives, genetic counselors may remain unprepared to effectively utilize AI tools, limiting both their own confidence in the technology and its potential to enhance patient care.

### Implications

Most genetic counselors are not using clinical genetics chatbots, and adoption is unlikely to accelerate unless professionals can trust that these tools provide accurate, reliable, and ethically sound information to patients. Participant responses reflect that chatbots could reduce the administrative burden for genetic counselors, though they express concerns about the chatbots’ ability to navigate complex genetic information and provide emotional support. Incorporation of clinical genetics chatbots will involve balancing AI implementation with patient-centered care, as participants noted concerns about the loss of personal connection in patient interactions. The fear that chatbots could replace genetic counselors underscores the importance of involving genetic counselors in conversations about using clinical genetics chatbots. As such, genetic counselors can guide chatbot involvement in the clinical space to ensure these tools complement rather than compromise the quality of genetic counseling.

Currently, the genetic counseling profession has few recommendations or formal guidelines on using clinical genetics chatbots for patient care. Findings from this study highlight the need for professional organizations to establish evidence-based recommendations that balance technological advancements with the concerns and considerations noted by participants, ensuring the technology supports but does not replace the human expertise and connection at the core of genetic counseling.

### Limitations

While this study had valuable findings, it has several limitations to interpretation. All participants were genetic counselors or prospective genetic counselors; therefore, the findings are most representative of this group and may not fully reflect the perspectives of other genetics professionals. Additionally, the sample size was limited when evaluating the experiences of individuals who have used clinical genetic chatbots in practice. This limitation made it difficult to draw meaningful conclusions about the firsthand experiences of those who have interacted with this technology in real-world settings. The geographical scope of the study was confined to North America, with certain states being more highly represented than others, and some were not represented at all. This uneven distribution of participants poses challenges in ensuring a diverse range of perspectives and experiences. Additionally, the lack of international representation makes it difficult to generalize the findings on a global scale, as healthcare systems, regulatory environments, and technological adoption vary significantly across different regions. Several biases may have influenced the study results. Because the data was based on subjective perceptions rather than objective measures of chatbot performance, participants’ responses may not accurately reflect the chatbots’ actual capabilities and their uses. Additionally, those who chose to complete the survey may have had a preexisting interest in genetic chatbots, which could have skewed the results positively. Conversely, a bias against chatbot integration may also exist, as genetic counselors and those training in the field may be hesitant to acknowledge the possibility of automation replacing aspects of their role.

### Future Directions

Future research should prioritize patient-centered research examining comprehension, trust, and satisfaction with AI-based tools in genetic counseling, as this study focused on provider perspectives. Understanding how patients perceive chatbot interactions is crucial to assessing their overall effectiveness and acceptability. Further investigation into institutional barriers is warranted, as many respondents identified their institution as a major obstacle, highlighting the need to explore administrative concerns such as cost, ethical and privacy considerations, and potential impacts on quality of care. Comparative research analyzing different chatbot platforms could also help identify which models are most effective for specific clinical tasks.

## Conclusion

This study offers valuable insights into the integration of AI-based chatbots in genetic counseling, presenting the current uptake, confidence, and potential benefits and challenges of this emerging technology. By alleviating administrative burdens, chatbots have the potential to enhance the efficiency of genetic counseling services, enabling GCs to focus on more complex, patient-specific responsibilities. This, in turn, could lead to improved patient access, reduced wait times, and lower rates of GC burnout. However, this study’s findings demonstrate that while chatbots show promise in streamlining administrative and routine clinical tasks, they have relatively low levels of use by GCs, rooted in lack of trust and integration difficulties. It is clear that genetic counselors and trainees are hesitant about employing AI-based clinical genetics chatbots, pointing to the need for standardized guidelines, training, and continued evaluation of chatbot effectiveness. Ultimately, this research contributes to the growing body of evidence on AI integration in healthcare. It underscores the importance of striking a balance between technological efficiency and personalized care in genetic counseling.

## Supporting information

NIH Coversheet

## Data Availability Statement

The survey can be found at: https://github.com/rlwaikel/AI_Clinical_Genetics. Raw survey data are available upon written request.

## Acknowledgements and Funding Information

This research was supported by the Intramural Research Program of the National Human Genome Research Institute of the National Institutes of Health and Bay Path University Genetic Counseling Program, and used the computational resources of the National Institutes of Health High-Performance Computing Services Biowulf cluster. The contributions of the NIH author(s) were made as part of their official duties as NIH federal employees, are in compliance with agency policy requirements, and are considered Works of the United States Government. However, the findings and conclusions presented in this paper are those of the author(s) and do not necessarily reflect the views of the NIH or the U.S. Department of Health and Human Services.

## Ethics Declaration

The Bay Path University Institutional Review Board deemed this study exempt on September 8, 2024. No identifiable data was collected. R.L.W. had full access to all the data in the study and takes responsibility for the integrity of the data and the accuracy of the data analysis.

