## Supplemental Tables for "Artificial Intelligence-Based Chatbots in Genetic Counseling Practice: Current Uptake, Utilization, and Perspectives"

| **Supplemental Table 1** |  |
| --- | --- |
| *Participant Demographics* |  |
| Genetic counselors' Current Area of Practice | Other: Immunology, operations/administrative, education, pharmacogenetics, lysosomal storage diseases, metabolic, hematology, preconception |
| Distribution of Institutions Among Genetic Counselors | Other: Government medical center, public or private university, industry, private practice, research institute, pharmaceutical industry |
| Distribution of Current Positions for Genetic Counselors | Other: Administrative, program director/educator |

| **Supplemental Table 2**  *General Chat Uses Outside of Clinical Practice* |
| --- |

|  | |  | Frequency | Percent |
| --- | --- | --- | --- | --- |
|  | General questions | | 128 | 77.1 |
|  | Composing text | | 101 | 60.8 |
|  | Entertainment | | 62 | 37.3 |
|  | Technology support | | 30 | 18.1 |
|  | Health related questions | | 21 | 12.7 |
|  | Composing computer code | | 10 | 6.0 |
|  | Help with child's homework | | 9 | 5.4 |
|  | Other (see below) | | 19 | 11.4 |
|  | Only use in clinic, no outside use | | 5 | 3.0 |
|  | *Total general chatbot users* | | **166** | **—** |

|  | Other: Online banking questions, hobbies, travel plans, home improvement project questions, design ideas, generating images, help with research, help with studying, scheduling, recipes, workout routines, summarizing |
| --- | --- |

**Supplemental Table 3**

*Self-Reported Knowledge of General Chatbots in Healthcare*

|  |  | Frequency | Percent |
| --- | --- | --- | --- |
| Genetic Counselors | Little to no knowledge | 95 | 60.9 |
|  | Intermediate knowledge | 59 | 37.8 |
|  | Highly knowledgeable | 2 | 1.3 |
|  | Total | **156** | **100** |
| Students | Little to no knowledge | 26 | 48.1 |
|  | Intermediate knowledge | 23 | 42.6 |
|  | Highly knowledgeable | 5 | 9.3 |
|  | Total | **54** | **100** |

**Supplemental Table 4**

*Uptake and Uses of Clinical Genetics Chatbot by Genetic Counselors (GC) and Students*

|  |  | | GC | Students |
| --- | --- | --- | --- | --- |
|  | Used | | 12 | 0 |
|  | Recommended | | 0 | 2 |
|  | Used and recommended | | 4 | 0 |
|  | Not used nor recommended | | 135 | 49 |
|  | Unsure | | 2 | 0 |
|  | *Total* | | **153** | **51** |
| Other uses of clinical chatbots: Research for positive results, tested but never implemented | | | | |

**Supplemental Table 5**

*Reported reasons for genetic counselors not using clinical genetics chatbots*

|  |  | | Frequency | | Percent |
| --- | --- | --- | --- | --- | --- |
| Technology not available | | 56 | | 41.5 | |
| Doesn’t work with current clinic workflow | | 56 | | 41.5 | |
| Healthcare facility limits the use of outside technologies | | 44 | | 32.6 | |
| Technology is not reliable | | 38 | | 28.1 | |
| No need for this technology | | 32 | | 23.7 | |
| Previous experience using a chatbot | | 4 | | 3.0 | |
| Other (see below) | | 20 | | 14.8 | |
| Unsure | | 16 | | 11.9 | |
| *Total participants (genetic counselors)* | | **135** | | **—** | |
| Other: Discomfort with technology, not currently seeing patients, concern for patient ease of use, distrust of technology, maintaining human connection | | | | | |

| ***Supplemental Table 6***  Respondents' Opinion of Patient Comprehension of Genetic Information When Provided by a Clinical Genetics Chatbot vs. a Human Counselor | | | |
| --- | --- | --- | --- |
|  |  | Frequency | Percent |
| Respondents' Opinion | Comprehension better with a chatbot | 3 | 1.5 |
|  | No difference in comprehension | 19 | 9.5 |
|  | Comprehension better with a human counselor | 145 | 72.9 |
|  | Unsure | 32 | 16.1 |
|  | *Total* | **199** | **100** |

| **Supplemental Table 7** |  |
| --- | --- |
| *Perceived Benefits of Clinical Genetics Chatbots for Providers and Patients* | |
| Providers | Other: No benefits, VUS reclassification notification, note writing, increase in patient education/consent in the absence of a GC |
| Patients | Other: No benefits, more efficient appointments, resource for review, lower wait times |

| **Supplemental Table 8** |  |
| --- | --- |
| *Participants’ Concerns About Offering Clinical Genetics Chatbots to Patients* | |
|  | Other: Adaptability to unpredictable circumstances/questions, ease of use for patients, assessing emotional state of patients, patient health literacy assessment, legal concerns, quality of customer service, missing important family history information, losing human interaction |

| **Supplemental Table 9** |  |
| --- | --- |
| *Training Experiences and Needs for Clinical Genetics Chatbots* | |
| Type of Training Received on Using Clinical Genetics Chatbots in a Healthcare Setting | Other: Through job position, through research study |
| Suggestions on Specific Training for Incorporating Clinical Genetics Chatbots into Clinic Workflow | Other: Training for non-genetics referring providers, training on chatbot strengths and limitations, how to integrate into practice, best practices for use, identifying/mitigating risks |

**Supplemental Table 10**

*Optional Free Response Answers from Participants*

***In what ways could clinical genetics chatbots be improved to meet your clinical needs?***

I would only use clinical genetics chatbots for scheduling appointments. There is no other instance in which I see it beneficial to use chatbots.

I think if I was given references for where the information the chat bot got its information from or what algorithm it used I would feel more confident.

have not used, so not sure. am a bit hesitant to use in my practice

It could definitely help with pre-test counseling.

In cancer there are many misconceptions that individuals have regarding genetics information and they can be poor historians where they may report a family history of ovarian cancer but with more prodding they actually switch to a cervical cancer diagnosis. I have no experience with chat bots but they may not be asking follow up questions to family history. Also, there is no emotional support a chatbot can provide to an individual.

I would want to spend time interacting with one so I could ensure that it works and replies to questions properly before using it with patients. I would also want to ensure that if a chatbot is being used, that a patient understands that there is a human they could reach out to with any questions as well.

I would want to have more control over the exact delivery of information than I have seen, for example, what a patient is told when they use GIA.

They would need to be on par with a human counselor in terms of their ability to elicit nuanced information - not just 'do you have a family history of X disease' but asking follow-up questions about patients' responses when they are vague, or catching on to common patient responses that indicate that they have misunderstood the question (for example, a patient answers yes when asked if they have a relative with ovarian cancer, but on further questioning it turns out it was most likely cervical cancer). Or a patient says they have a family history of type 2 diabetes but upon further questioning the history is more likely to reflect type 1. In addition, the chatbots would need to be able to respond to unique patient needs and adapt to their individual situations - ie I don't think it would be helpful for a chatbot to provide management recommendations when these would be general and not patient-centered nor would take into account what has already been done, what extenuating circumstances there might be etc. I am not sure a chatbot will ever be able to be on par with human counselors in these ways.

continued education efforts to quickly find information

Allow us to deliver services to more patients, focus direct patient care on the patients that need us most.

Include metrics or options for patients who do not finish the chatbot questionnaire. A way to say they are exiting the chatbot early and why so that we as clinicians know if information is missing or there were gaps of education using the chatbot.

Field FAQs to help triage the more complex questions requiring a genetic counselor to the genetic counselor

I would need more oversight from an actual GC checking all responses.

It would be amazing if there could be a way to facilitate the implementation of clinical genetic chatbots into the EMR of any hospital. Or maybe there could be a certification by a medical organization that could prove the chatbot's reliability.

review a condition's features with a family who had an abnormal newborn screen for that condition

I feel they would me more helpful with scheduling, billing questions, or for negative results if they are adapted correctly. I think they would also be helpful for general patient questions online that are very common to save time.

Family history collection could be beneficial though I would still need to go back and ask clarifying questions. I think it could be very useful in notifying at risk family members as letters are old fashioned and easy to lose!

I have no idea what they are currently doing so no idea how they can be improved.

Genetics specific "training"

Currently, my experience with AI is negative owing to inaccurate dispersal of information from ChatGPT, Google search AI, etc. and therefore I would not expect an AI system to be accurate.

More responsive/interactive

I think there needs to be improvement on their ability to counsel about a positive or complicated VUS result. Psychosocial counseling is important for these results and that is hard to achieve with a ChatBot.

I just don't trust them so maybe if they were verified by real genetic counselors. But also I'm putting in time to learn how to deliver information and I don't want a chatbot doing it for me

I would most certainly want to ensure that they are only receiving information from the most up-to-date guidelines and resources. Especially in cancer genetics, the guidelines can change frequently, so I would be concerned about a chatbot neglecting to include updated screening information if it was informing a patient of their risks with a positive mutation.

Help disclose negative results and answer any questions related

scheduling

I work primarily with rare/ultra-rare diagnoses in children, and lots of VUS results from WES, WGS and CMA. Often, very little is even published about these conditions/variants, so I would likely not trust that a chatbot is adequately trained to locate relevant information for my use in patient care. I think an appropriately trained chatbot could be useful for education of patients/families about basic genetics and genetic testing principles, e.g. prior to or following a clinic or telehealth visit. I also think that for specific indications, it could be helpful for screening/triage/directing referrals, though not necessarily for our typical patients who have a high level of complexity of referral indications.

They can't.

pre-test counseling and getting family history

The could be helpful to gather very general, basic information prior to an appointment so more time can be spent asking higher level questions during the appointment.

If the bot feature allows for more natural form of verbal communication then it would be able to meet my clinical needs.

HIPAA compliant

Easier to integrate into clinical care. There are many institutional hurdles to being allowed to use these types of tools for clinical care.

In general, how confident are you in the ability of clinical genetics chatbots for the following purposes. I don't know how to answer this question if I have not used it.

I'm not sure; perhaps drafting documentation for return of results

Usage of clinical genetics chatbots can help patients understand general genetic information in a comprehensive way

I think it could help with the daily tasks that do not need critical thinking (e.g. some scheduling, basic education etc)

I would love to be able to use chatbots at our institution for things like help with triaging referrals, consults, helping non-genetics providers order genetic testing, helping patients understand insurance pre-authorization / cost, family testing, etc. We just don't have the technology at our institution.

Identify patients who need a referral, give general genetics information, collect family history

Unsure. It would be nice to use it to make clinic templates and handouts based on indications that I could check, but my institution doesn't offer this so I am not sure how to try it out.

We need to acknowledge that chatbots in the GC field must be highly vetted and programmed in order to achieve clinical utility, which is more easily applied to locations like individuals clinics. In a laboratory setting, where lots of different institutions use the same laboratory but in customized ways, any chatbot solution would be further complicated in its design due to the variation and complexity of different user group needs

More information about scope/utility outside patient facing practice ie medical education, generation of differentials, creating LMN/patient letters, finding organizations & supports for patients etc

Make it easier to adjust them, make them cheaper, make them more accessible, teach healthcare providers how to build them and use them

Increased access to these technologies

Time. I would need time to verify that the chatbot is accurate (e.g. collating accurate history info from EMR)

I think ultimately humans and chatbots need to work alongside to provide quality care. If chatbots could take a more first pass approach and keep information confidential, I may consider using it.

writing letters, findings resources, summarizing data or research.

My clinic has not yet attempted to incorporate chatbots routinely. I do think it can save time in certain capacities, as is already seen with other electronic forms on communication. However, I am not sure how they can be improved because we have not tried them yet. I would need to trial.

This is an area that I am beginning to conduct research in as well, in reviewing current literature it appears that AI struggles with nuanced understanding of ever rigid clinical guidelines, has a tendency to use outdated guidelines for recommendations, and also gives inconsistent answers to the same questions being asked (if not beholden to predefined answers). While I think that some aspects of information gathering and patient education can be useful currently, the three aforementioned areas of difficulties need to be addressed prior to its widespread use in clinic. Recent litigation in Colorado against a hospital's genetics team as well as a genetic testing company, alleging for wrongful life and birth, leads me to believe that similar litigation could be opened against systems that employ AI if incomplete or inconsistent recommendations are provided.

literature searches for providers

I think there is potential for the technology to be used for documentation/charting/letter creation.

more data

Be more trustworthy

A chatbot could potentially be used in the place of an FTE GCA to provide general info or release simple test results

unsure

unsure; would need to try them first!

Have better safe guarding and data protection

Comprehensive chart review and summary equivalent to what I do now, comparable family history

In the tools I have tried, there have been way too many inaccuracies for me to even trust it if using it myself

screening to refer patients

For patients who need point of care genetic testing (ie pancreatic cancer patients), chatbots could be used to provide pre-test education at an oncology appointment and save the patient a trip to genetics (unless they are positive)

I require more training before I am comfortable using them

Obtaining family histories prior to session; disclosing normal results in some settings

Unless it earns a degree, performs requisite clinical rotations, and passes a board exam, I will not trust a chatbot as I would trust myself or a colleague. To that end, I'm not sure it can be improved to help me

expansion into tasks needed in research genetic counseling (research consenting, disclosure of research testing)

get family history in advance, get patient history in advance, explain negative/normal results

I've had poor experiences trying to get medical info from chatbots/AI. They would have to get more accurate and have a way to send patients to a real person if needed.

Summarize HPI from other specialties for clinic prep, screening for info that could be relevant to Genetics evaluation.

I think Chatbots could allow patients who are fairly confident they want genetic testing to have the ability to have testing/counseling without needing to see us, giving us more time and shorter waits for patients who have more questions or concerns.

***What if any issues have you encountered when using clinical genetics chatbots, if any?***

I would need a government-approved, patient facing chatbot where I could pre-screen virtually all possible responses to make sure they weren't providing misinformation.

Timing of our reach with chatbot, should this be connected to an appointment? How to maximize engagement with the chatbot?

Lack of free response/open-ended options for patients to input their own answers or explanations when the questionnaire has fixed options

The lack of psychosocial counseling

The company deploying the chatbot didn't have a great interface with our EMR, so patients would think they were getting a spam text/email and not engage, or complain to our hospital

They only provide very basic information and often go in circles when a patient asks a question.

institutional support and regulation, low uptake of offers to a genetic counseling appointment via chatbot

Expensive, lack of ability to edit, some patients don't like them, some patients prefer them so much that they don't want to talk to a provider, not always accurate for a patient's unique condition or context

When using GIA to screen for patients who should have genetic testing, patients who met criteria were missed by GIA.

***Please use this space for any additional comments or suggestions related to using chatbots in genetic counseling.***

patient's limited accessibility and access to chatbots

Incorrect information concerning genetics concepts, recurrence risks, etc.

Although there are algorithms we use for following a workflow in genetics, each patient has different needs and expectations. It seems like it would be near impossible to program a chat bot to anticipate all of the needs of a patient in order to give accurate information, or to successively relieve a patient's concerns

I am looking forward to the future of chatbots but I think it will be important to keep them tightly regulated. I think there are some places where human genetic counselors are better, and places that chatbots can take over some aspects and make things more efficient.

Training should include how genetic counselors can continuously review and edit the information provided by these chatbots to patients.

I think chatbots would be a great addition to genetic counseling workflows. However, I worry about accessibility and that this will lead to larger healthcare disparities.

I worry about a chatbots ability to provide the "counseling" aspect of genetic counseling. A chatbot could not have real and meaningful empathy or an understanding of real-world issues (language barriers, financial barriers, access barriers, etc.), which I think would negatively impact its ability to do its job and provide information in a meaningful manner. A chatbot might be more useful in a financially privileged institution, but I'm not sure how helpful it would be in safety net hospitals.

If a chatbot is going to be used in genetic counseling practice, it is important to understand what the patient experience is from start to finish to best communicate about the chatbot.

with all the "scandals" involving chatbot hallucinations/flat out incorrect information I don't have any level of trust in this technology at this point.

I worry they they would replace GCs and yet not be accurate, or not offer emotional support, and then what if the patient just checked out and did not listen and there is no way to check their understanding.

I think their use would need to be backed up by a human available to triage any challenges that arise while using the chat bot, similar to customer service situations that now use chatbots and transfer you to a real human if they cannot help you.

It's going to be used in some way, but GCs/Genetics professionals need to be involved and guide how, when, and to what extent such technology is integrated in our profession. This isn't something to be viewed as an inevitable revolution that we just have to 'get ready for'/view as having no agency in shaping.

I think a benefit of human genetic counselors is having the ability to adapt our education to the specific patient. A ChatBot can only be programmed to get so personalized and I would anticipate they'd be pretty generic. My education can look drastically different between appointments depending on the needs of the patient and I don't see that kind of adaptability being possible with this kind of technology. I'm not closed off to the idea but I would need to see it in practice.

Chatbots have a lot of bias that is currently unexplored. AI is detrimental to the environment. AI has inaccurate understanding of data, especially in current models. It's been shown that asking AI how many "R"s are there in the word ___ is inaccurate, so I can imagine it is simply not ready for clinical use.

I am an Asst. Director of a genetic counseling training program and we do discuss this with our students and provide examples.

I think chatbots can be promising aids to genetic counselors in the future. I think my biggest fear is that other providers who order their own genetic testing without consulting or referring to a GC may rely too heavily on the chat bots to disclose results, discuss screenings, etc.. I feel as if chatbots can be used as a tool and not a replacement, they could be very advantageous. Any outcome where the chatbot is replacing the work of a GC could have troubling implications if not monitored properly. I work in a state that has some of the biggest education barriers in the country, and I have had patients that required multiple different and simplified explanations well below the 6th grade learning level that I’m not sure chat bots could consistently detect and adjust to meet those needs.

As someone who works in a clinic with a 6+ month long wait list, i think we as providers have to set aside some of our fear and discomfort and embrace technology that can help reduce wait times and get more patients the services they need

I use chatbots a lot in my educational role and none of the questions asked captured that non-clinical role.

I think that people should be considering the cost of chatbots in comparison to genetic counselors. Not only financial but also their environmental and social cost/impact ([<https://earth.org/environmental-impact-chatgpt/>](https://earth.org/environmental-impact-chatgpt/))..

I think there is likely a space for them; but I would hope we use technology to offload more administrative tasks and therefore allow clinicians more time with patients. I think its integration is likely to be affected by how the field of GC moves forward and chooses to define itself and vice versa.

I think that it could be helpful, but I could see increased patient frustration if their exact questions aren't being answered. Additionally, I would like to see how it discusses uncertain information. We have a lot of patients with VUS' and sometimes we aren't sure how to follow-up or explain results so it would be hard to teach AI how to respond.

I think AI is a powerful tool to support counselors in the non-patient facing aspects of work (ie gathering differentials, formatting LMN, suggesting support organizations), but many of the nuances of human conversation are required for results disclosure, family history intake, etc. I see more utility in chat bots answering patients questions like “what is the cost/coverage?” or “what is this test?” rather than interpretation of results or intake of family history

I think that these tools could be more helpful for the tasks that do not need as much human connection such as family history or general education. I am apprehensive about their use in regards to the aspects of genetic counseling that require more counseling and attending to our patients' emotional needs. I would be afraid of GCs getting too reliant on these technologies and losing the parts about our field that make us special.

I am a little weary of chatbots being used. For one, I am not sure how well they would be able to personalize care for a patient.

I suspect there's a place for these tools but wouldn't want to eliminate the human touch for helping people navigate complex info and their feelings around testing/results

If implementing a chatbot it would be essential for it to be accurate, but also to know how to screen for patients who would really benefit from 1:1 discussion

I have high hopes that chatbots could actually increase the human aspect of our profession because genetic counselors could focus on counseling and values based decision making. My biggest concern is that counselors won't have the motivation or training to do the counseling side of our job. If that happens I believe that GCs will become obsolete, because the information gathering and giving part of our job can be done just as well and less expensively by a chatbot.

Understanding it's history in other related fields

I think there is also an ethical consideration for how chatbot/AI information is obtained, and the amount of power that it uses, that people don't give consideration to.

I think it would be useful for (some) negative results and carrier screening mostly, but otherwise I don't think it would be helpful
