## Supplemental Figures for "Artificial Intelligence-Based Chatbots in Genetic Counseling Practice: Current Uptake, Utilization, and Perspectives"

### Slide 1
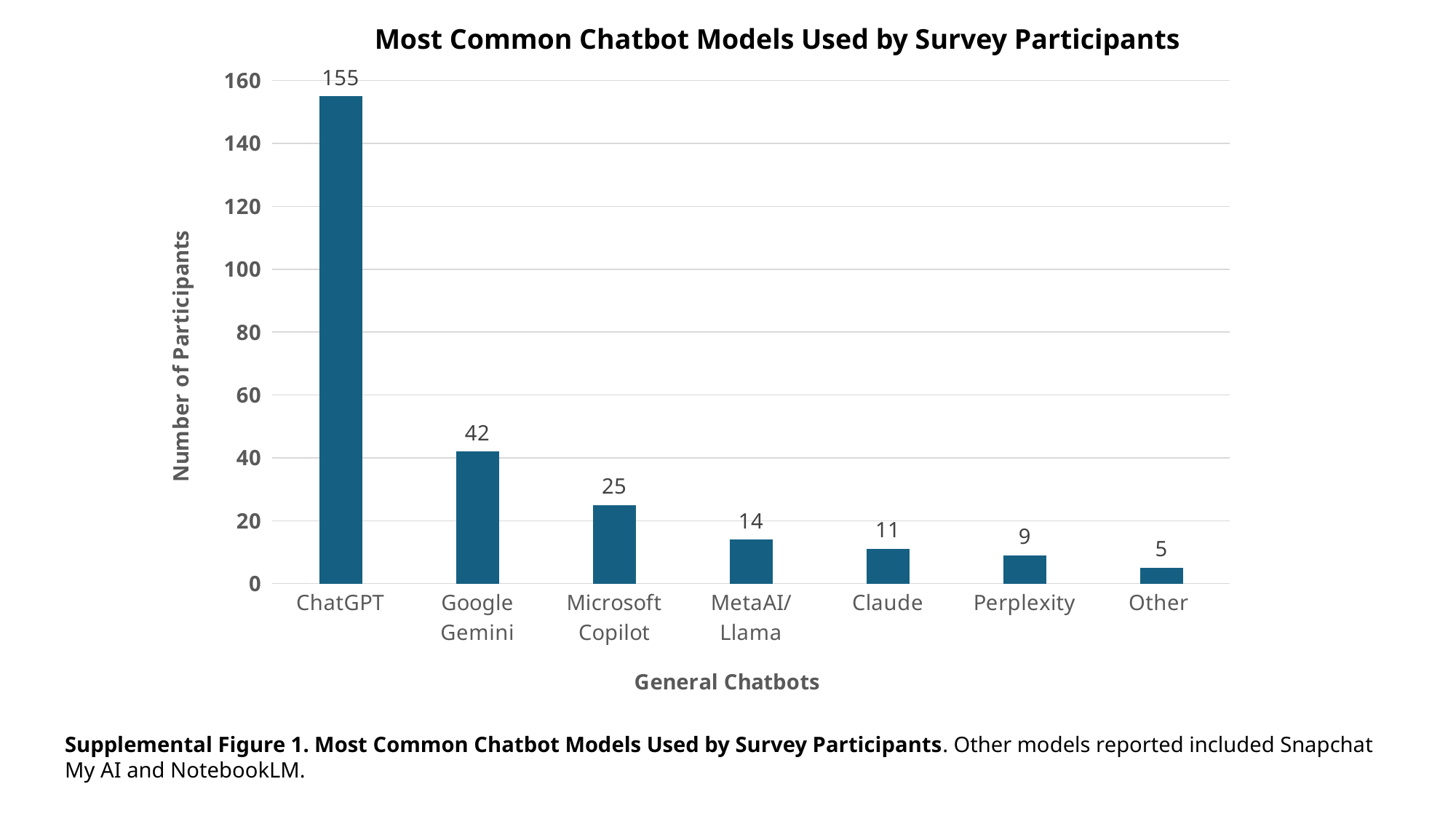

Most Common Chatbot Models Used by Survey Participants
#### Chart
| Category | |
|---|---|
| ChatGPT | 155.0 |
| Google Gemini | 42.0 |
| Microsoft Copilot | 25.0 |
| MetaAI/Llama | 14.0 |
| Claude | 11.0 |
| Perplexity | 9.0 |
| Other | 5.0 |Supplemental Figure 1. Most Common Chatbot Models Used by Survey Participants. Other models reported included Snapchat My AI and NotebookLM.

### Slide 2
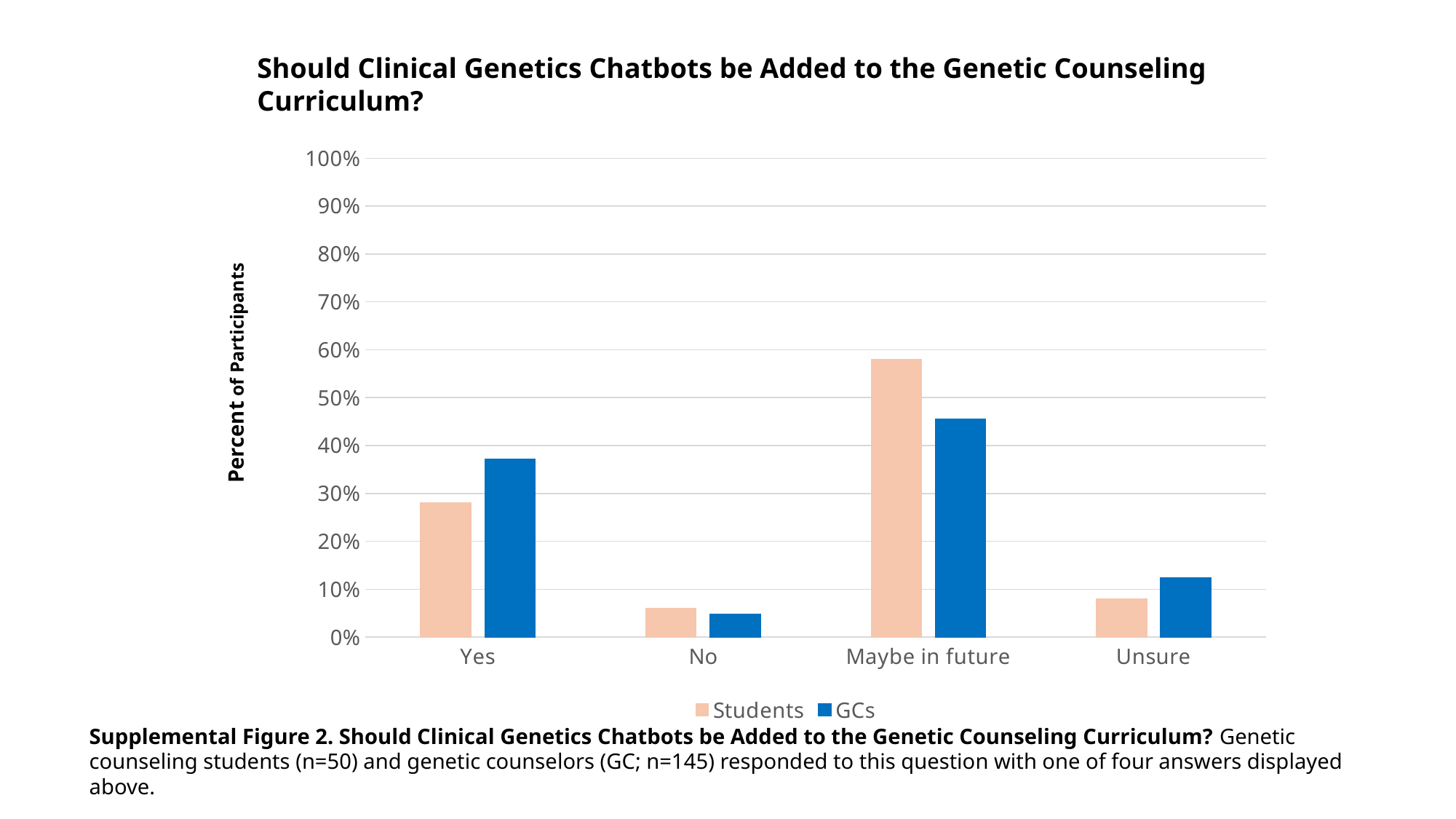

Should Clinical Genetics Chatbots be Added to the Genetic Counseling Curriculum?
#### Chart
| Category | Students | GCs |
|---|---|---|
| Yes | 0.28 | 0.372 |
| No | 0.06 | 0.048 |
| Maybe in future | 0.58 | 0.456 |
| Unsure | 0.08 | 0.124 |Percent of Participants
Supplemental Figure 2. Should Clinical Genetics Chatbots be Added to the Genetic Counseling Curriculum? Genetic counseling students (n=50) and genetic counselors (GC; n=145) responded to this question with one of four answers displayed above.
